# Capacity building in the global polio eradication initiative: individual wins and institutional opportunities, a qualitative analysis

**DOI:** 10.64898/2026.02.10.26346031

**Authors:** Koumudi Thanda, Anna Kalbarczyk, Oluwaseun Akinyemi, Humayra Binte Anwar, Wakgari Deressa, Eric Mafuta, Yodi Mahendradhata, Piyusha Majumdar, Ahmad Omid Rahimi, Olakunle Alonge

## Abstract

The Global Polio Eradication Initiative (GPEI) is known to be one of the most successful disease eradication initiatives with massive reductions in polio transmission as well as its contributions to strengthening health systems and capacity building. Existing analyses provide limited insight into how capacity was developed, which forms of capacity were strengthened or neglected, and how these efforts were experienced by implementers. Understanding these factors can be helpful as GPEI accelerates transition and integration into national health systems. This study examines GPEI’s capacity-building efforts from the perspectives of GPEI workers across multiple roles, levels and countries.

We conducted a qualitative secondary analysis of semi-structured key informant interviews with global, national, and subnational stakeholders involved in GPEI. Data were coded inductively and deductively analyzed through Capacity Building Pyramid by Potter and Brough 2004. Country-based co-authors reviewed findings for contextual accuracy.

We found that GPEI substantially strengthened individual and infrastructure capacity through large-scale training of frontline health workers, development of immunization infrastructure, and establishment of operational. Polio-funded platforms enabled outreach to hard-to-reach populations and were leveraged for other health priorities. Despite substantial investments, capacity building has remained largely individual training and program centric. There was insufficient integration into national systems leading to constrained country ownership and, lack of diverse trainings leading to limited supervisory and management capacity.

Lessons of capacity building done under GPEI provide important guidance for future planning, and implementation of large-scale interventions. These findings highlight the need to move beyond predominantly individual-focused training toward deliberate, systems-level capacity strengthening. As polio transition planning accelerates, global health policymakers and planners must prioritize systems-level capacity strengthening, sustainable financing, and national governance to ensure continuity of immunization and public health functions beyond GPEI. Leveraging GPEI assets for broader health system strengthening can accelerate progress toward universal immunization and health care.

## Introduction

The Global Polio Eradication Initiative (GPEI) was launched in 1988 by the World Health Assembly with the ambitious goal of eradicating polio by 2000. Inspired by the success of smallpox eradication, the initiative sought to leverage mass immunization campaigns, surveillance systems, and global coordination to eliminate poliovirus. Despite significant progress, reducing polio cases by 99%, full eradication has yet to be achieved, with endemic wild poliovirus in Pakistan and Afghanistan and variant polio in 46 other countries (1).

GPEI operates as a public-private partnership led by the World Health Organization, UNICEF, Rotary International, the U.S. Centers for Disease Control and Prevention, and national governments, with additional contributions from the Bill & Melinda Gates Foundation, Gavi, and local partners. The initiative historically followed a largely vertical approach, prioritizing disease-specific strategies such as polio vaccinations only, targeted campaigns, and surveillance activities. While this approach has been highly effective in reducing polio cases globally, it has also been criticized for creating parallel systems that do not always integrate well with existing health infrastructures (2). Recognizing this as well as reduced need for active surveillance, GPEI recently shifted its focus toward integrating its activities within country-led immunization programs, emphasizing management and coordination support for broader immunization efforts (3,4).

Capacity building in global health programs refers to the efforts to strengthen the capabilities of individuals, organizations, and systems to effectively implement and sustain health initiatives (5,6). Historically, many capacity-building efforts have focused primarily on training individuals, often through short-term programs or external technical assistance, rather than addressing systemic and structural barriers such as workforce development, leadership, and governance frameworks. This sometimes resulted in fragmented improvements and limited long-term impact. In recent years, there has been a growing focus on standardizing the concept of capacity building and establishing best practices for monitoring, evaluation, and learning (7). Effective capacity building is increasingly recognized as a practical and sustainable approach to strengthening health systems and as an ethical obligation for global health partners, requiring equitable partnerships and mutual learning (8). However, many global health capacity-building efforts perpetuate neo-colonial models, where experts from high-income countries (HICs) provide short-term support in low- and middle-income countries (LMICs), potentially without sufficient engagement of local stakeholders or attention to sustainability, leading to limited systemic change and reinforcing existing power imbalances (5,8,9). Sustainable and impactful capacity building thus requires long-term partnerships, local leadership, and a shift towards models that prioritize institutional strengthening and shared governance (6,10,11).

Capacity building has been a core element of GPEI for many years, and its approaches have varied across different contexts and levels of the health system (12–14). The initiative has leveraged a range of strategies, including the use of geographic information systems for real-time surveillance and response, surge capacity support, trainings, and systems strengthening to address the challenges of polio eradication. These efforts have been instrumental in improving surveillance of polio cases and outbreak response, immunization coverage, and health system resilience, although they have also faced challenges such as funding constraints and integration with local health systems (15,16). Furthermore, GPEI has built a wide range of assets and large global workforce (17,18). However, as GPEI concentrates on transitioning out of countries and toward integration with national health systems, an examination of its capacity-building efforts is crucial. Program integration in a health system relies on leveraging existing capacities and infrastructures to maximize efficiency, and ensure sustainability (19–21). Existing capacities such as trained personnel, data systems, and organizational structures provide a foundation for the introduction of new programs, reducing the need for additional resources, and facilitating smoother and more rapid implementation. This approach, along with new capacity building efforts, is particularly beneficial in resource-constrained settings where maximizing the use of available resources is crucial. This dual strategy of leveraging existing capacities and investing in targeted capacity building is the best practice for achieving effective, sustainable program integration in diverse health system contexts (22,23). Existing analyses highlight the overall benefits of capacity built under GPEI but provide limited insight on how capacity was developed, what types of capacity were strengthened and/or overlooked, and the perspectives of GPEI workers through all this. This study aims to address these gaps by examining GPEI’s capacity-building efforts from the perspective of implementers and stakeholders across multiple countries, identifying how the initiative had an impact on skills, tools, and systems at different levels of a health system.

This research is part of STRIPE (Synthesis and Translation of Research and Innovation in Polio Eradication) which is a collaborative consortium between Johns Hopkins Bloomberg School of Public Health (BSPH) and seven institutional partners representing areas where GPEI activities occurred. STRIPE is a mixed methods study that applies an implementation science lens to consider the actors, processes and contexts for program activities within the GPEI to systematically identify and translate the lessons learned. These findings aim to inform and improve policy, planning, program design and implementation of eradication and other large-scale public health interventions moving forward (24).

## Methods

The seven institutions of the STRIPE consortium were selected from focus countries that represent each epidemiological classification for polio (endemic, outbreak, at-risk and polio-free) in 2017, different geographical regions, income classifications, conflict situations, and countries that could serve as influential leaders within their region on disease eradication (24). *Table 1* describes the selected countries and institutions.

**Table 1.**
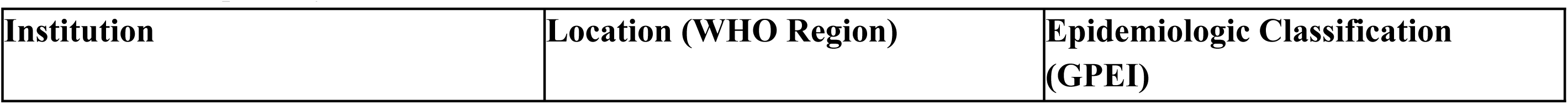
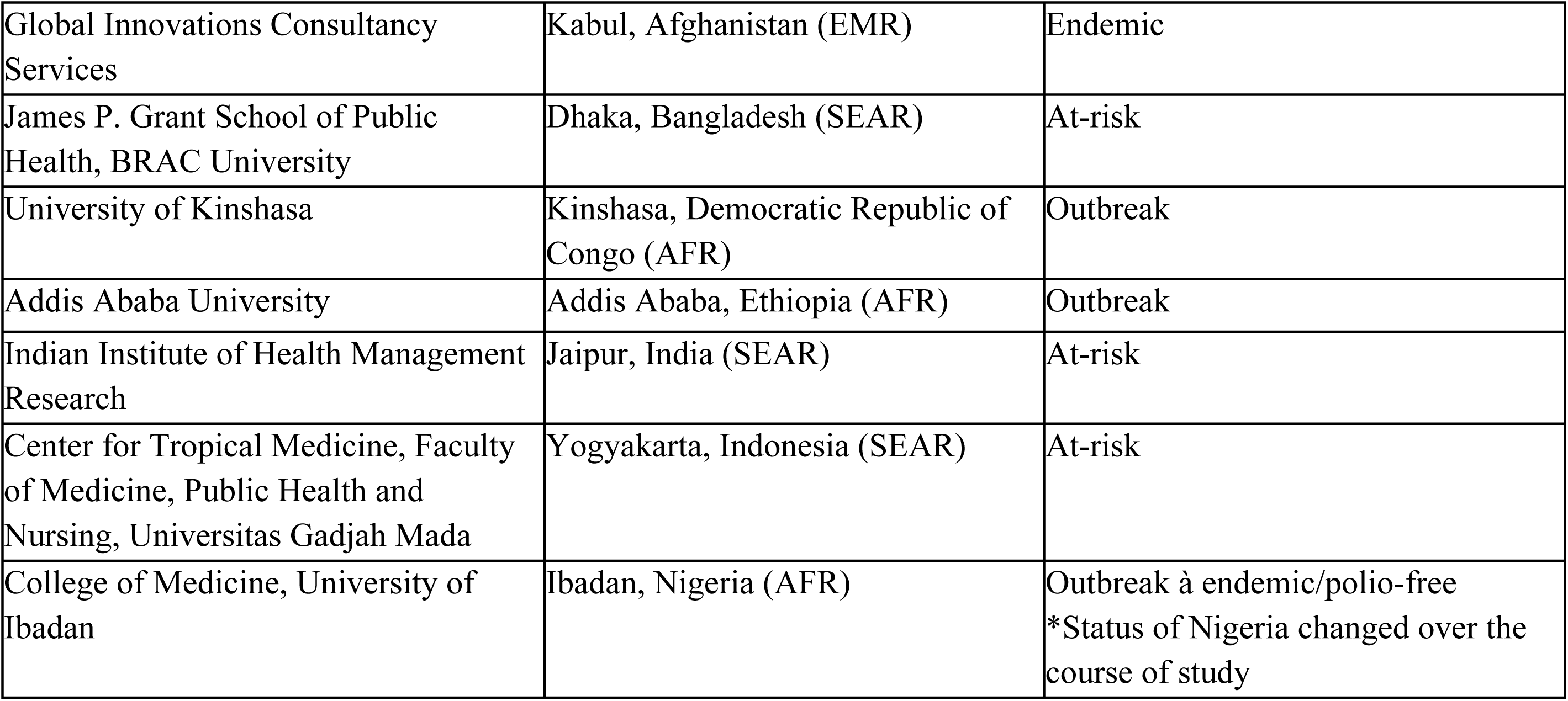
Description of selected countries and institutions in STRIPE.

The STRIPE project encompasses four key phases: knowledge mapping, synthesis, packaging, and dissemination and uptake. The first year of this collaboration focused on the knowledge mapping activities, utilizing a sequential explanatory mixed methods design including a large-scale quantitative global survey, and key informant interviews (KIIs) (25,26).

Respondents, all members of the polio universe, were national-, sub-national-, and frontline actors defined as any individual 18 years of age or older who worked on some level of polio eradication activities for 12 or more continuous months between 1988 and 2019, identified from a quantitative survey (24,27). KIIs were conducted from a nested sample of survey respondents with the goal to further explore implementation challenges of polio eradication activities, addressing these challenges, and their intended and unintended outcomes. The nested sample was identified through a review of survey responses for respondents who could speak to challenges faced during polio eradication activities across domains (i.e. health system levels, geography, organizations, and expertise) and were representative of overall survey.

The KII interview guide prioritized questions on polio organization and change over time, contextual challenges, strategies, including capacity-building activities, utilized to address challenges faced, and key lessons learned. The KIIs were face to face and in-depth, semi-structured interviews, conducted by two experienced interviewers at the global level and 3-4 experienced, trained interviewers in each focus country. A training manual was developed to ensure standard interview processes (i.e., recruitment processes, transcription, memo drafting, and data management) were met and country-level interviewers were trained in a 2 or 3-day institutionally determined training. Prior to participation in the study, each study participant provided informed consent after they were given the study purpose and the voluntary nature of participation in plain language. The recruitment and data collection for the main study was done between August 2018 - December 2019. The interviews were conducted in participant’s preferred language (English or local languages) between February and May 2019. Each interview took 70-100 minutes, recorded and then transcribed, translated into English for analysis. A codebook was developed according to two implementation science frameworks - Consolidated Framework for Implementation Research and the Socio-Ecological Model - to capture key lessons learned (28,29).

A total of 194 interviews were conducted in all focus countries as well as at the global level. Data management, coding and analysis were conducted in Dedoose, a qualitative data analysis software. Four reviewers conducted a pilot test on two interviews from different countries, applying the codebook to both interviews. Analytics were run in Dedoose to detect discrepancies between reviewers, and a meeting was held to review these issues and reach consensus. Post-analysis data validation was conducted, including cross-checking key findings with interviewees.

For this paper, the coded KIIs were searched for key text terms such as “capacity”, “training”, “human resources”, “learning” and “self-efficacy”. Existing codes included “organization”, “individual”, “individual characteristics”, “lessons learned”, “strategy”, “challenge”, “successful” and “self-efficacy”. This process yielded 50 excerpted responses from various KIIs: 13 from respondents in Ethiopia, 13 from Afghanistan, 12 from global stakeholders, 9 from India, 1 from Indonesia and 1 from Bangladesh. These excerpts were then re-coded using the analytical framework for Capacity Building described by Potter and Brough (30). Of the identified excerpt pool, 28 interviews contained relevant information and were retained for the final analysis.

The secondary coding was conducted by lead author with experience in global health research and health systems qualitative coding, who systematically reviewed each excerpt and assigned it to one or more categories within the Potter & Brough framework (e.g., tools, skills, staff, systems). The coder used a deductive approach: each excerpt was read multiple times, initial codes were applied using the framework categories, and coding decisions were documented to ensure transparency. Coding was periodically discussed with the second author to ensure proper understanding with the information and framework.

### The Analytical Framework

To address issues in standardizing capacity building, Potter and Brough (2004) identified a hierarchy of needs specific to capacity building activities, providing leaders with a logical framework for planning, implementing and evaluating interventions (30). They found that regardless of the intentions or skill of individuals, systemic and organizational issues continuously impact the ability of the individuals to create lasting change. The authors identified four interconnected levels of capacity building, from base to tip: structures, systems and roles; staff and infrastructure; skills; and tools. Nine separate but interdependent component types of systemic capacity building feed into these levels: performance, personal, workload, supervisory, facility, support service, systems, structural, and role capacity. These two groups can then be organized into a logical hierarchy and inform how success or failure at any of these levels influences the others (*Fig 1*).

**Fig. 1:**
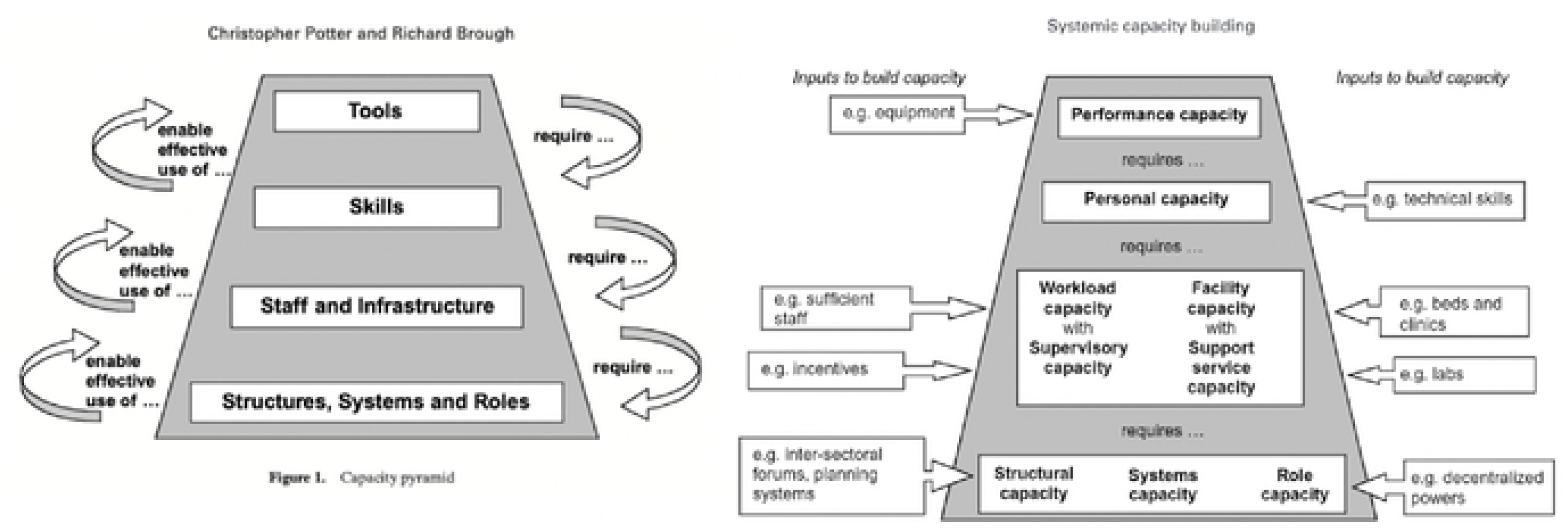
Potter and Brough Pyramid of Effective Systemic Capacity Building (30). Alt text: Two flat topped pyramid diagrams illustrating the ‘Pyramid of Effective Capacity Building.’ One on the right is the overall pyramid with four horizontal layers stacked from bottom to top, with arrow leading up to next levels with text “enable effective use of…” to show the hierarchical relationship between different forms of capacity. There are also arrows going from top of pyramid to the bottom with text “require…”. The base layer holds structures, systems, and roles. The second layer represents staff and infrastructure. The third layer represents skills. The fourth layer represents tools. The second flat topped pyramid completements the first with definitions, and examples of inputs to build these capacities. The base layer is made of structural capacity (e.g. inter sectoral forums, planning systems), systems capacity, and role capacity (e.g. decentralized powers). Second layer up has workload capacity (e.g. sufficient staff) with supervisory capacity (e.g. incentives), facility capacity (e.g. beds and clinics) with support service capacity (e.g. labs). Third layer up has personal capacity (e.g. technical skills). And last layer has performance capacity (e.g. equipment).

This research was submitted to the Johns Hopkins Bloomberg School of Public Health Institutional Review Board and deemed to be “non-human subjects research.” Additional approvals were obtained from: Institutional Review Board of the Ministry of Public Health, Afghanistan, the James P. Grant School of Public Health Ethical Review Committee, the Kinshasa School of Public Health Institutional Review Board, the Institutional Review Board of the College of Health Sciences of Addis Ababa University, the Institutional Review Board for Protection of Human Subjects, IIHMR, the Medical and Health Research Ethics Committee, Gadjah Mada University, and the National Human Research Ethics Committee, Nigeria. Informed consent was obtained from all research participants. Survey participants were provided with a written consent statement prior to accessing the survey. Oral consent was obtained from all participants.

## Results

### Participants

The results are based on the subsample of 28 unique KIIs relevant to capacity building efforts extracted from 194 KIIs conducted as part of the STRIPE study. The 194 KIIs were nested in a survey of 3,659 respondents including participants from all seven countries – and the results of the full survey and KIIs are published elsewhere (24). The 28 unique KIIs include respondents from global, national, and sub-national levels across India (5), Ethiopia (8), Afghanistan (6), Bangladesh (1), Indonesia (1), and global stakeholders (7). It includes individuals with expertise in various GPEI programmatic activities (e.g., advocacy, communications, and surveillance), representing different entities (e.g., national and subnational government entities, implementing organizations, donors and frontline health workers). *Table 2* displays the demographic characteristics of all KII participants and the subsample extracted for the capacity building analysis; gender was not collected on the surveys to ensure privacy for participants but was noted upon interview if that individual was selected for a KII (27).

**Table 2.** Characteristics of Respondents.

|  | **STRIPE KIIs (N=194)**<br>**n(%)** | **Final Sample (N=28)**<br>**n(%)** |
| --- | --- | --- |
| Global | 18 (9.3) | 7 (25.0) |
| Afghanistan | 28 (14.4) | 6 (21.4) |
| Bangladesh | 17 (8.8) | 1 (3.6) |
| DRC | 23 (11.9) | - |
| Ethiopia | 30 (15.4) | 8 (28.6) |
| India | 25 (12.9) | 5 (17.8) |
| Indonesia | 26 (13.4) | 1 (3.6) |
| Nigeria | 27 (13.9) | - |
| Global | 18 (9.3) | 7 (25.0) |
| National | 84 (43.3) | 13 (46.4) |
| Sub-national | 70 (36.1) | 7 (25.0) |
| Front line | 22 (11.3) | 1 (3.6) |
| GPEI partners | 61 (31.4) | 12 (42.8) |
| Government | 96 (49.5) | 10 (35.7) |
| Implementing organizations | 23 (11.9) | 4 (14.3) |
| Academic organizations | 5 (2.6) | 2 (7.1) |
| Other | 9 (4.6) | - |
| Male | 144 (74.5) | 24 (85.7) |
| Female | 50 (25.5) | 4 (14.3) |

### Tools

Capacity building activities in the GPEI were largely concentrated at the individual levels of the capacity building pyramid (i.e. skills and tools), through trainings on polio disease, vaccine administration, interacting with caregivers, compiling reports, marking houses, transportation, reporting (31). The trainings primarily targeted supervisors, frontline health workers and key community members, to provide skills on vaccinating children (31). In the capacity building pyramid, the highest level is tools, pertaining to the availability of essential resources like tools, equipment, and finances which enable workers to effectively carry out their jobs.

Key informants (KI*s*) widely acknowledged that GPEI investments played a crucial role in increasing the availability of essential tools and resources for effective vaccination efforts. Funding supported the procurement of supplies including vaccines, cold chain equipment, vaccine carriers, and vehicles, many of which remained within national health systems and strengthened broader immunization programs. They shared that the tools and resources provided through GPEI investments often extended beyond polio, supporting other health initiatives by contributing logistic capacity and trained personnel.

> *“The polio program had a lot of funds and supported other programs by providing vaccine carrier and kerosene” (Ethiopia National GPEI Partner 05)*

> *“The resources and the infrastructure for bringing new cold chain, vaccine carriers and other things which remained with the system, which can be utilized for other things” (India National GPEI Partner 13)*

KIs also pointed to gaps in the available tools that hindered their performance. Outdated technology, inconsistent vaccine supply, and financial constraints limited the effectiveness of their eradication efforts. In some cases, countries struggled to integrate emerging technologies that could enhance surveillance and immunization tracking, while others faced ongoing shortages of vaccines, slowing progress toward eradication.

> *“Still countries are lagging behind, and I do not understand because [polio] should be given at the same times as the [routine immunizations] …it is not yet up to the level that it should be to be able to ensure that everyone has enough vaccine … for the eradication” (Global GPEI Partner 04)*

KIs shared GPEI investments overall had a positive impact on the provision of tools as noted by the vaccine availability, cold chain infrastructure, and logistical support, benefiting broader health efforts. All of this improved the performance capacity of the frontline workers. However, gaps in technology, inconsistent supply, and financial constraints were limits to their capacity.

### Skills

The skills level represents the second layer of the capacity building pyramid and focuses on personal capacity. Respondents shared that GPEI investments focused on enhancing the personal capacity of health workers - strengthening their knowledge, technical abilities, and confidence to execute polio immunization effectively. Across multiple countries, respondents highlighted the success of training programs in equipping frontline workers, program managers, and field epidemiologists with essential immunization competencies, ranging from community mobilization to disease surveillance and micro-planning.

> *“Polio brings the capacity development of those frontline workers who were not aware about what is mobilization or what is communication or talking to mother. We now have better-informed frontline workers who have better capacity of communicating and mobilizing people.” (India National GPEI Partner 13)*

> *“[GPEI] built the skill of all leaders that participated in the program. If you go to the sub-city, woredas [districts], they all know microplanning.” (Ethiopia Frontline Government Worker 02)*

> *“[GPEI] started field epidemiology training for provincial polio officers. This will help detect the trends and respond in time for any indication of polio. This would curb polio transmission. This is a good thing in capacity building of provincial officers and EPI managers which never happened before.” (Afghanistan National Government Worker 10)*

In Ethiopia and Afghanistan, KIs credited GPEI’s structured, long-term training programs for strengthening the technical competencies, delivery, and overall implementation ability of the health workforce. They shared that intensive training initiatives increased the pool of skilled personnel and led to creation of new positions that supported program implementation. However, these trainings were centered on polio logistics and vaccine delivery, with comparatively little attention given to interpersonal skills, patient communication, and community engagement. KIs mentioned that training often excluded workers considered non-essential for specific skills, even though they could have benefited from the knowledge. This was particularly true for workers coming from non-health sectors who lacked knowledge of how to communicate with patients.

> *“When we provide training, we give only to the focal person or coordinators instead of providing for those un- relevant personnel which causes the challenges from the capacity issues. In past, the director or the health managers had participated on the training who did not work directly on the program rather than giving directives” (Ethiopia Subnational Government Worker 05)*

> *“Our HR, maybe the focus is too, too focused. The capacity building or training was too focused on technical matters, immunization, vaccine delivery. Even though communication skills were also very important. In my opinion it was still lacking, very lacking.” (Indonesia National Government Worker 10)*

> *“We have seminars and workshops for campaigns. I would suggest they could be increased. This is good for the capacity building of staff as many of our staff come from non-health sector. They are not nurses or midwives. It is good to conduct workshops for people working in the program especially the team leaders even if they are midwives or nurse.” (Afghanistan Subnational Government Worker 03)*

### Staff and Infrastructure

While the overall number of skilled workers improved, KIs shared that supervision and monitoring systems did not grow accordingly. Some KIs raised concerns around accountability, corruption, and frequent staff rotation without transition protocols under GPEI.

> *“There is a high staff turnover at facilities… there is a gap in hand over, they don’t handover and orient the substitute” (Ethiopia Frontline Government Worker 02)*

> *“Sometimes some people make problems, because they are not professional, and they don’t have capacity; they are here on personal relations. They cannot perform job and achieve targets because they are not sound technically.” (Afghanistan National Government Worker 01)*

In terms of physical infrastructure, KIs consistently highlighted the role of GPEI in improving facilities and logistics. GPEI left behind facilities that continue to be useful for other health programs. The program’s logistics, surveillance systems, and vehicles have also been repurposed for other vaccine-preventable disease surveillance and immunization efforts.

> *“The human resources and the network can be used by anyone. People can use their capacity of human resources. People can use our facilities; we have enough facilities in every province. We have also supported clinics in other service provision from our resources through UNICEF and CDC.” (Afghanistan Subnational Government Worker 05)*

> *“From the positive aspect, it’s impossible to quantify but I think that there have been countless, numerable really examples of how polio resources, assets, staff, systems, logistics have benefited other public health initiatives…support that all of the polio resources have provided to vaccine preventable disease surveillance which is really mostly measles surveillance, lab supported measles surveillance as well as the other vaccination campaigns that take place by using the polio assets whether they be the vehicles or the staff to help plan and implement and even evaluate and asses the quality of these campaigns.” (Global GPEI Partner 04)*

However, infrastructure related to biomedical research or domestic vaccine manufacturing was underdeveloped. One KI from India mentioned the need to produce vaccines domestically which would help sustain immunization programs without external reliance and maintain a polio free status.

> *“Now more emphasis will be to daily maintain the polio free status overemphasis will be to really indicate our professional commitment switching over from Trivalent to Bivalent vaccine and introduction of IPV then you know….those are the important areas and then being self-sufficient financially to meet the requirement of the program and then we have to have really build our own capacity to produce vaccine.” (India National GPEI Partner 08)*

### Structures, Systems and Roles

The final layer of the P&B Model is structures, systems and roles (SSR) focuses on how and if the mechanisms of the health system and its structures enable workers to apply their skills and function. Within SSR, the system capacity examines the capacity in efficient flow of information, finances, and managerial decisions, while structural capacity analyses decision-making forums, intersectoral collaboration, and accountability processes. The KIs highlighted how the investments by GPEI led to notable improvements in the SSR capacities in their settings. Many also described missed opportunities for sustainability.

### Structures and Systems Capacity

The KIs shared that the GPEI played a large role in introducing components of their health systems that were previously unestablished like accountability frameworks, microplanning, cold chain processes, immunization delivery, and developing a whole workforce on health programming. And in countries like India, GPEI was able to generate new data and serve previously underserved areas.

> *“The structure polio has created—like the review mechanism at the block level, district level, state level, and the periodicity and intensity—now provides a forum for reviewing and progressing any other public health program.” (India National GPEI Partner 13)*

Key informants acknowledged that while the GPEI was viewed as a vertical program, in practice, they said it significantly supported the integration of immunization and health system activities across multiple countries. As mentioned before, GPEI provided infrastructure, tools, and workforce capacity that was leveraged by other programs.

> *“I think to the credit of polio program it has helped to build [the] system. People used to say it was a vertical program and [that it] destroy[ed] systems. I don’t think [so]—we strongly believe that it actually helped to build [systems].” (India National GPEI Partner 14)*

KIs described how GPEI resources were adapted for other health goals, particularly in resource limited areas. In Bangladesh, the national EPI structure and surveillance systems for diseases like rubella were strengthened through polio resources. In Ethiopia, other programs like child health, screening, malaria and nutrition were often integrated with GPEI program. An informant from Afghanistan emphasized that the facilities and human resources developed through GPEI remained accessible and useful across the health system. And in India, new systems were introduced to underserved regions.

> *“Maternal neonatal tetanus elimination efforts, measles mortality reduction efforts, yellow fever, the response to outbreaks of Ebola and expand it and talk about the work in emergencies that often happens and it’s polio staff are there, they’re located at the sub-national level so they can help to participate in the organization of a responsive to a cholera outbreak or to tsunami response.” (Global GPEI Partner 07)*

GPEI’s emphasis on community involvement was notably important in Ethiopia. Communities gained awareness about polio and developed a sense of ownership to proactively identify and report cases. In addition, collaborations with regional and zonal health departments helped strengthen the capacity of new health workers.

> *“The polio program is not only implemented by the health worker but also the community members, so a lot of training was done. Now, if you go to any regions, everyone knows about polio and the immunization program. In the polio eradication program, every house is knocked at, and the community knows what polio vaccination means.” (Ethiopia National GPEI Partner 05)*

> *“During the past, you only checked for suspected cases from those patients who came to the health facilities, but now, when the community-based approach comes, a member of the community is the one to search for the cases inside their own community.” (Ethiopia Subnational Implementing Partner 04)*

KIs also shared that GPEI fostered collaboration among various global stakeholders and in extending health services to underserved areas. The coordination between GPEI and multilateral organizations like UNICEF and CDC facilitated the expansion of healthcare services beyond immunization, particularly in Afghanistan.

KIs then noted shortcomings of GPEI. The massive resources and infrastructure developed for polio eradication were not always leveraged to strengthen routine immunization programs or broader health initiatives. Some described GPEI as narrowly focused, filling gaps in fragile health systems creating an unsustainable dependency. And while polio infrastructure was established and valuable, the assessment of its impact was rarely documented.

> *“If a similar level of support and resources had been provided to strengthening routine immunization, we would probably—well they would say—we would be further along in doing that or any other health initiative where the same level of resources could have been implicated.” (Afghanistan National Government Worker 10)*

> *“I think the program unfortunately hasn’t done a really good job of documenting [infrastructure and assets] so there are vignettes, there’s anecdotes, some of it is documented of course, but really understanding the impact of the assets on the ground has been difficult.” (Global GPEI Partner 04)*

Another major challenge identified by KIs was that GPEI did not sufficiently build local capacity for countries to take full and sustained ownership of immunization programs. They shared those countries facing economic, political, and social instability relied on the external funding and technical support and as GPEI shifted focus or transitioned out, the routine health services weakened.

> *“We can be criticized of the polio program for not having supported in a way that was building more country ownership, more domestic engagement, domestic funding, human resource capacity at the country level… this is probably a handful of countries or maybe two hands of countries—countries which don’t have polio as their own program. They have so many other economic problems, wellness problems, political problems, unrest problems that polio is just the reviewing agent of a much more profound weakness in these countries.” (Global GPEI Partner 03)*

KIs from Ethiopia shared that the increased demand for cold chain management and vaccine stock maintenance during polio campaigns at times created logistical challenges for other immunization programs.

> *“During campaign times, there were lots of stocks and it created pressure. It had pressure because you had lots of stocks and you must manage the cold chain very well; there might be stock outs, so you have to take care of that. During that time, it had an effect on other vaccinations because your focus was on the campaign. It is a must to conduct routine immunizations, but it is affected by the campaign.” (Ethiopia Frontline Government Worker 02)*

### Roles capacity

Respondents emphasized GPEI’s focus on improving personal capacity, in turn improved the rapid decision-making and implementation at various levels in a health system. The mass capacity building in individuals allowed former GPEI workers to use their skills in other public health programs within their country and globally.

> *“Now almost every staff of the government knows about the program and their capacity developed now they can take their own initiative own decision. so now already the capacity development is happening …not only at the government level… most of the other organizations you will find there are some people who work for the polio program… the capacity was not building here, but this group of people also actually support all over the country…all over the world.” (Bangladesh National GPEI Partner 12)*

Though there was success in increasing roles capacity through GPEI, KIs pointed to power imbalances, decision making authority, and ownership of data, especially between local governments and GPEI workers. Program workers held more power in the collected data, limiting the local government authority and ownership of their country’s data.

> *“A Nigerian minister said, “You have been doing this in our country for decades and yet I don’t have any data managers in my ministry who can actually do the kinds of analyses you do, and you’re supposed to be building our capacity, what’s wrong?” And it was just such a telling moment because it was like-- he was basically saying, “I would like to own the data, but I can’t and by nature of your presence, you’re kind of tying our hands and we have to listen to what you’re saying about what this data says.” (Global GPEI Partner 08)*

As GPEI’s funding decreases in countries and begins to transition out of countries, KIs shared that local governments will be forced to make essential decisions with limited information and expertise. Global respondents shared that the complex funding structure for GPEI can be another challenge for the countries. With the limited focus on building roles capacity within each system and creating a plan for sustained programs, KIs warned of panic among countries.

> *“As polio budgets start to decline, countries, as well as the polio partners and countries, are going to have to figure out how to right-size their support.” (Global GPEI Partner 07)*

> *“GPI, like I said doesn’t give money to governments, it gives it to agencies, I hope and I believe that the agencies need to be a lot smarter about how they first identify the technical minimums that need to be on the ground because that is their job and number two, make the hard decisions about cutting their workforce in big ways and actually building government capacity instead of filling government capacity.” (Global GPEI Partner 08)*

### Contextual Variation

The reported GPEI’s capacity-building activities varied across countries, largely reflecting differences in sociopolitical, economic, and geographic contexts. For example, KIs from Afghanistan mentioned conflicts and security concerns, which hindered implementation and sustainability of the program.

> *“Security is making problems for us in selection of staff, in training and capacity building it is also affecting program implementation and monitoring of field activities. So we can say if we had security we would have selected competent people according to the qualification for the implementation of program. So we would have been able to select qualified people without interference from government people or the opposition groups and that would have enabled us to implement quality program” (Afghanistan Subnational Government Worker 06)*

KIs from Ethiopia noted that they worked within the political context of the country and highlighted the role of community-based volunteers like the Health Development Army (HDA) who play a central role in immunization and service delivery. However, the decentralized structure posed unique challenges for capacity building and program monitoring. As one KI explained, the large number of low-literacy volunteers made effective supervision difficult.

> *“The health development army is one part of the health system. However, they can’t read and write…it is difficult to monitor them because they are several in numbers. For example, there might be 30-100 who are in 1 for 5 teams in one “kebele” which is the lowest administrative unit. This is very difficult to follow up which makes the monitoring very poor. But if there are 5-6 trained volunteer health workers or HDA within one lowest administrative unit it might improve the monitoring system. So since they are several in numbers it becomes difficult to monitor and bring the expected outcome.” (Ethiopia National GPEI Partner 03)*

In contrast, certain countries described having stronger baseline public workforce capacity which enabled GPEI to build upon existing systems. Alternatively, in other settings GPEI might have created parallel systems placing additional strain on local health workers and raising concerns about burnout and sustainability.

> *“In PAHO, you had a higher level of public workforce capacity to begin with so that really helped but it begs the question, should we focus on that workforce capacity versus basically fly people in…the burnout comes from needing to work around the system versus through it…would it have been better to go slower and more diligently versus try to just smash this thing through? …like if you don’t have the basic systems that you would need already on the ground, maybe you consider not doing it until you do have the systems.” (Global GPEI Partner 08)*

A few respondents spoke positively of GPEI’s role in introducing critical infrastructure and processes while others were critical. For example, KIIs from India, Afghanistan, and Bangladesh, credited the polio program with creating new systems for surveillance, microplanning, cold chain management, and workforce expansion. They also mentioned that these components later supported other health initiatives in the country. Conversely, in other settings, particularly Ethiopia, respondents were more critical of GPEI’s impact. These informants raised concerns about overreliance on external actors, corruption, funding and unsustainable structures left behind during the GPEI transition. Global actors echoed the critiques.

> *“A large number of data managers and officers are reduced from their jobs so the work loads are transferred to government institutions but in the government institution there is no skilled and experienced human resource so to manage this all it needs capacity building. The capacitated body left the institution for their own reasons.” (Ethiopia National Academic Organization Worker 03)*

## Discussion

This study analyzed GPEI’s contribution to capacity building across individual, organizational, and systemic levels. Findings show that GPEI achieved notable success in developing individual capacities and tools for immunization, yet more systemic gaps like ownership remain.

One of GPEI’s most highlighted capacity building activities was its substantial investment in the long-term technical training of health workers, including vaccinators, community health workers (CHWs), cold chain managers, among others. Informants shared that many of these trained workers transitioned into broader health programs and emergency response within their countries, supporting maternal and child health, measles campaigns, and outbreak control. These results are consistent with existing literature on GPEI trained workers’ impact in countries (12,32–34). Additional data found that density of CHWs in LMICs has been shown to improve polio coverage more than the density of nurses and doctors (35) and that the presence of CHWs increases immunization coverage in LMICs (36). So this improvement in staff capacity led by GPEI in our findings, could have contributed to improved immunization and health care coverage in LMICs over the years.

The development of health workforce is particularly important in the context of persistent global health workforce shortages (37,38), now projected to reach 10 million by 2030, decreased by a previous estimate of 18 million (39). These shortages are often linked to countries’ limited capacity to finance, train, remunerate health workers, and to provide adequate resources (40). GPEI contributed to addressing these gaps by supporting increases in health worker density through financing and resource provision, especially in underserved areas. Beyond expanding workforce numbers, ensuring these workers are adequately trained and equipped with the right skill mix is also critical. Training to build a skilled workforce is a fundamental aspect of the Capacity Building Pyramid, as individual-centered trainings enable health workers to effectively use tools and implement health programs (30). However, to most effectively learn and use these skills, adequate staff and infrastructure are necessary, including an appropriate skill mix to workers, establishing reporting and monitoring systems (30). Our findings indicate that while GPEI’s training efforts are widely valued and many GPEI-trained staff have successfully moved into leadership or technical roles within national or regional health programs, and have been called upon to support other outbreak responses (18,33). This demonstrates the significance contribution of GPEI investments to strengthening the broader health workforce and supporting priority health programs beyond polio (33).

Yet we found there remains a demand for diverse trainings with broader skill sets and GPEI critiqued for its limited reach, often excluding lower-level or non-technical staff. GPEI trainings were also criticized for strong emphasis on technical skills which often came at the expense of developing interpersonal and supervisory skills like communication, leadership, and community engagement. These interpersonal skills are essential for global health workers for effective service delivery, leadership, team coordination, and sustaining community trust (41,42). For CHWs in particular, these skills are critical for understanding cultural expectations, fostering engagement, and serving as a bridge between communities and the health system (42). Without these competencies, health workers may struggle to manage community interactions, communicate effectively, or lead programs. Including interpersonal skills in training would improve patient outcomes and enhance community participation (41).

Diversifying trainings would also create a mix of skills among workers and enable efficient task shifting, further strengthening workers’ capacity. A well-balanced skill mix enables individual workers to take on multiple roles, reducing the need for separate workers for specific task and lowering operational and training costs (43). This flexibility allows for effective task shifting, which involves delegating tasks across different health worker cadres to optimize limited human resources. Task-shifting can be inefficient if not well planned and has concerns about quality of services and systemic weaknesses (44,45). However, it has been a strategy in low-resource settings to address workforce shortages (45). If efficiently planned, task shifting can save costs and build an even stronger health workforce which ensures quality of care (44,45). This would also reduce reliance on reactive task shifting, improve cost savings for immunization activities, minimize bottlenecks in service delivery, and improve the overall health programs (45,46). Additionally, achieving this requires broader and more inclusive training programs that equip workers with the competencies needed to confidently take on diverse roles (45,46). The demand for trainings of diverse skillset from frontline GPEI can improve the individual capacity of workers as well as the systemic capacity and retain the massive health workforce built by GPEI.

GPEI’s contributions extended beyond workforce development, encompassing substantial investments in infrastructure, tools, and processes that supported and strengthened broader public health programming. The literature aligns with our findings, showing that the initiative introduced and improved critical assets like cold chain systems, vehicles, data management systems, laboratories, and microplanning frameworks. Many of these tools were retained or repurposed for other health programs, including maternal and child health services, deworming campaigns, COVID-19 response, and outbreak control (47,48). For example, surveillance and laboratory infrastructure supported by GPEI supported disease surveillance and response strategies in the WHO African region, which led to detection and management of multiple communicable diseases beyond polio (49).

According to the frameworks like Capacity Building Pyramid, staff- and infrastructure-level capacities enable the effective use of individual skills, but long-term sustainability and comprehensive system improvement require strong systems, structures, and clearly defined roles (30). The legacy of GPEI demonstrates that while physical and technical assets are vital, their full impact is realized when properly embedded with the health system that can adapt and sustain these improvements overtime (15). Our findings show that weak documentation, outdated technologies, and limited supervisory capacity pose threats to the long-term sustainability of these tools and infrastructure. Informants mentioned concerns about countries’ dependency on external decision-making, funding, and no formal integration of processes into national systems, which threaten sustainability of the program. According to the Capacity Building Pyramid, these gaps indicate underdeveloped systems, structures, and roles capacity (30).

Research suggests that when decision-making is driven by external actors, countries may lack the institutional experience and autonomy required to take ownership and authority over decisions like workforce management, campaign planning, and resource allocation (50–52). This perpetuates dependency on external actors, which can weaken countries’ ability to manage immunization programs after donor transitions, unless power and control are deliberately shifted to local actors (53,54).

Informants also raised concerns about limited ownership over data and tools, which in some cases, such as in Nigeria, led to tensions between governments and external partners. This prompted policy responses like data protection mandates to reclaim authority over surveillance and immunization data (55). Such systems-level ownership is necessary for countries to sustain progress in polio eradication.

While gaps in ownership and systems integration pose challenges, informants also highlighted that GPEI fostered cross-sectoral and multi-level collaboration, particularly in hard-to-reach regions. These collaborations connected national and regional governments with global partners, aligning resources, technical expertise, and enabling public health responses across countries, a finding that is well supported by the literature (50,56). Such collaboration contributes to building the “systems, structures, and roles” capacity described in the Capacity Building Pyramid (30). These partnerships have been found to be key drivers of success post-transition in GPEI-priority countries and LMICs with past immunization programs, and can further strengthen evidence-based immunization programming (54,57–59). To sustain progress, it is essential that national governments and global partners uphold and leverage these collaborative platforms.

Afghanistan is the only country that remains polio endemic in this sub analysis. While many countries with GPEI-supported capacities are transitioning, Afghanistan’s focus remains on controlling and eradicating polio (60). Progress in the country has been repeatedly disrupted by conflict and political instability (61). The Taliban’s takeover in 2021 led to interruptions in international aid, withdrawal of critical global funding streams, and restrictions on health and humanitarian engagement (62). These systemic and political barriers threaten the sustainability of GPEI-trained health workers, tools and the infrastructure built over decades. Respondents from Afghanistan described limited government capacity, high turnover, and risks of corruption, issues that further complicate efforts to maintain essential polio functions. These further underscore the need for a uniquely tailored, context-sensitive approach for sustaining progress toward polio eradication in Afghanistan.

### Limitations

Firstly, findings are based on key informant interviews, which may be subject to social desirability and recall bias. Secondly, many respondents were closely affiliated with or benefited from GPEI, which could lead to optimism bias, where they may emphasize the positives and successes over gaps and limitations. Thirdly, the data used in this analysis is a sub search with specific terms, of the larger study and we might not have fully captured all mentions of capacity building in each interview. Similarly, these respondents also represent a small fraction of the large workforce that contributed to 40 years of GPEI programming. Though we were intentional to include diverse set of KIIs from all levels and regions of the program. Additionally, this subset has limited representation from countries such as Indonesia and Bangladesh. This introduces a geographic bias in the data and limits the generalizability and completeness of our findings which might overrepresent ideas and perspectives from countries with greater data presence. The evolving global immunization landscape, with changing priorities and funding structures, may limit the applicability of some findings. Lastly, the STRIPE project began before the Taliban takeover in 2021 and the drastic change in power and governance has uniquely impacted Afghanistan hence the findings may not fully reflect the current context. Despite these limitations, the study provides valuable insights into key capacity building successes and challenges, highlighting the importance of holistic training, inclusive governance, sustainable infrastructure, and local ownership to inform future immunization and public health policy and planning.

## Conclusion

Our findings emphasize the need to shift from individual-focused training to systems-level capacity strengthening, especially during the GPEI transition period. GPEI made significant contributions to health systems strengthening by building workforce capacity, developing critical infrastructure, and fostering multi-level collaboration. However, sustainability challenges persist, particularly around inclusive training, supervisory skills, local ownership, and integration of systems and processes. Addressing these gaps through comprehensive capacity building is essential for maintaining immunization successes and advancing broader global health goals. Lessons from GPEI offer valuable guidance for future global health policies and initiatives aiming to build resilient, adaptable, and locally owned health systems.

## Data Availability

Data can be available upon request.

## Acknowledgments

We acknowledge Alexandra (Lexy) Jamison for contribution to the conceptualization of this paper. We are grateful to all the members of STRIPE academic consortium for their support for this research activity. We also thank all the participants who participated in this study for providing their valuable time and insights. Finally, we give our special thanks to all the data collectors for their hard work in collecting data and contributions.

